# Targeted County-Level Interventions Achieve Epidemic Control Without Statewide Lockdowns

**DOI:** 10.1101/2025.10.15.25338091

**Authors:** Haridas K. Das, Tao Hu, Mauricio Santillana, Lucas M. Stolerman

**Affiliations:** Department of Mathematics, Oklahoma State University, Stillwater, OK, USA; Department of Mathematics, University of Dhaka, Dhaka, Bangladesh; Department of Geography, Oklahoma State University, Stillwater, OK, USA; Machine Intelligence Group for the Betterment of Health and the Environment, Network Science Institute, Northeastern University, Boston, MA, USA; Harvard T.H. Chan School of Public Health, Boston, MA, USA

**Keywords:** Epidemic modeling, Hotspots, Mobility data, Targeted interventions, Pandemic preparedness, Policy implementation

## Abstract

When a new pathogen emerges, public health authorities must act rapidly to mitigate its spread while minimizing socioeconomic disruption. Despite extensive debate on localized epidemic control, no model-based study has systematically evaluated county-level interventions for statewide epidemic suppression in the United States. We present a metapopulation model that integrates county-level mobility data to identify epidemic hotspots and assess targeted intervention strategies for pandemic preparedness. We identify epidemic hotspots as counties that generate disproportionately large statewide epidemics when serving as outbreak origins. These hotspots align with population-dense and highly connected locations but provide sharper spatial contrast than traditional centrality metrics. Targeted interventions reducing the basic reproduction number (*R*_0_) only at hotspots achieve substantial epidemic control—reducing statewide epidemics by 60–90% in four representative states (Oklahoma, New York, Florida, and California)—without requiring broad lockdown measures. Hybrid strategies combining moderate reductions in *R*_0_ (10–30%) with partial mobility restrictions (30–80%) from hotspot counties, while preserving activity elsewhere, achieve control equivalent to full suppression at hotspots. This framework demonstrates that strategic, location-specific interventions can replace blanket pandemic responses. It provides state and county decision-makers with a quantitative tool for prospective pandemic planning, enabling rapid hotspot identification and intervention design grounded in empirical mobility networks.

## Main

Effective pandemic planning requires identifying where interventions will have maximum impact while minimizing societal disruption. The COVID-19 pandemic exposed the need for rapid and strategic public health responses to emerging pathogens [1, 2]. While much has been learned, the challenge of designing control strategies that are both effective and minimally disruptive remains a central concern for policymakers. As we look toward future pandemics, there is a pressing need for interpretable models that can inform targeted interventions—particularly at the scale where real decisions are made. In the United States, this means state- and county-level modeling frameworks that support planning by local officials.

The challenge of identifying critical locations for epidemic control is fundamentally a network problem. Metapopulation models, which explicitly account for movement between spatial units [3, 4, 5], provide a natural framework to address this challenge by extending classical mechanistic models to incorporate the essential role of human mobility. However, traditional approaches to hotspot or risk-map identification often rely on network centrality measures, population density or flow thresholds, and historical surveillance data, thereby overlooking the dynamic interplay between local transmission intensity and network connectivity that determines a location’s true epidemic potential.

The COVID-19 pandemic catalyzed a wave of studies leveraging mobility data and metapopulation models to evaluate disease spread and interventions [6, 7, 8, 9, 10, 11, 12, 13, 14, 15, 16, 17, 18]. However, these efforts have primarily focused on rapid response to the COVID-19 pandemic, often tailored to model calibration, retrospective forecasting, or scenario analysis specific to that context—rather than on developing generalizable frameworks to guide response strategies for future outbreaks. While these studies demonstrated the value of mobility data for understanding epidemic dynamics, they provide limited guidance for prospective intervention planning where pathogen characteristics and optimal response strategies remain unknown.

Existing implementations of metapopulation models face several limitations for operational pandemic planning. First, many rely on theoretical mobility models (such as gravity models [19]) rather than empirical movement patterns. Second, most focus on large-scale dynamics rather than the county-level resolution, where many public health decisions are implemented in the United States. Third, few provide systematic methods for ranking locations by their potential to amplify epidemics, instead relying on ad hoc assessments of network structure or population size.

Here, we address these limitations by developing a framework that integrates population flows data from GPS-enabled mobile devices with a mechanistic metapopulation model to systematically identify epidemic hotspots and evaluate targeted intervention strategies. Our approach differs from previous work in three key aspects. First, we use comprehensive data from GPS-enabled mobile devices covering all U.S. counties to parameterize realistic movement networks rather than theoretical approximations. Second, we systematically identify hotspots by simulating outbreaks seeded in every county and measuring each location’s potential to amplify statewide transmission. Third, we demonstrate that targeted county-level interventions at hotspots can achieve statewide epidemic control without blanket responses. In particular, hybrid intervention strategies, combining moderate reductions in both local transmission and outbound mobility at hotspots, can achieve epidemic control equivalent to complete local suppression while reducing intervention burden.

Our computational framework provides a mechanistically transparent and interpretable tool for pandemic preparedness, allowing decision-makers to understand how changes in transmission or mobility impact epidemic outcomes. By leveraging pre-pandemic mobility patterns, our model enables rapid hotspot identification and intervention design at the geographic scale of public health decision-making. The framework thus enables more adaptive and practical policies, demonstrating that it is possible to improve outcomes without imposing overly aggressive interventions.

### Simulating Disease Spread With a Mobility-Informed Metapopulation Model

We developed a computational framework that enables pathogen-agnostic simulations for pandemic preparedness. Our metapopulation approach extends the classical SIR model [20, 21] by integrating SafeGraph county-level human mobility data between U.S. counties [22, 23]. To demonstrate the spatiotemporal detail of the mobility data and the model’s capabilities, we simulate epidemic dynamics across the network of 77 counties in Oklahoma (Fig. 1). Daily population flows between Payne and Oklahoma counties (Fig. 1a) were affected by COVID-19 stay-at-home orders, with a marked reduction in mobility around March 28, 2020 (shaded grey region) [24], illustrating how policy interventions can reshape mobility networks (Fig. 1b). We utilized pre-lockdown inter-county flows to compute time-dependent *flux matrices* Φ(*t*), comprising 77 *×* 77 entries per day and parameterizing our mobility-informed metapopulation model (see Methods). These matrices capture pathogen-independent connectivity patterns, enabling simulations designed for prospective pandemic planning. We simulated an epidemic seeded in Oklahoma County (Fig. 1c), which has the largest population in the state, to illustrate the role of mobility in spatial disease propagation. To represent a scenario of weak transmissibility, all counties were assigned a baseline transmission rate *β*_0_, set slightly above the recovery rate *γ*, except for Oklahoma County, where a seven-fold higher transmission rate was used to reflect elevated contact rates in a more urbanized area (i.e., *β*_seed_ = 7 *× β*_0_). This factor was chosen to generate a sufficiently large outbreak within the pre-lockdown simulation window. The simulation also included an initially infected population in the seeding county (*I*_*seed*_(0) = 3). Infections spread outward over time, driven by the underlying mobility network. Normalized prevalence time series for selected counties reveal substantial variability in local epidemic curves (Fig. 1d). State maps also exhibit variability in peak intensity and timing across counties (Fig. 1e). Notably, the western counties experienced higher and earlier peaks compared to the eastern part of the state, as a result of regional mobility connections.

**Figure 1:**
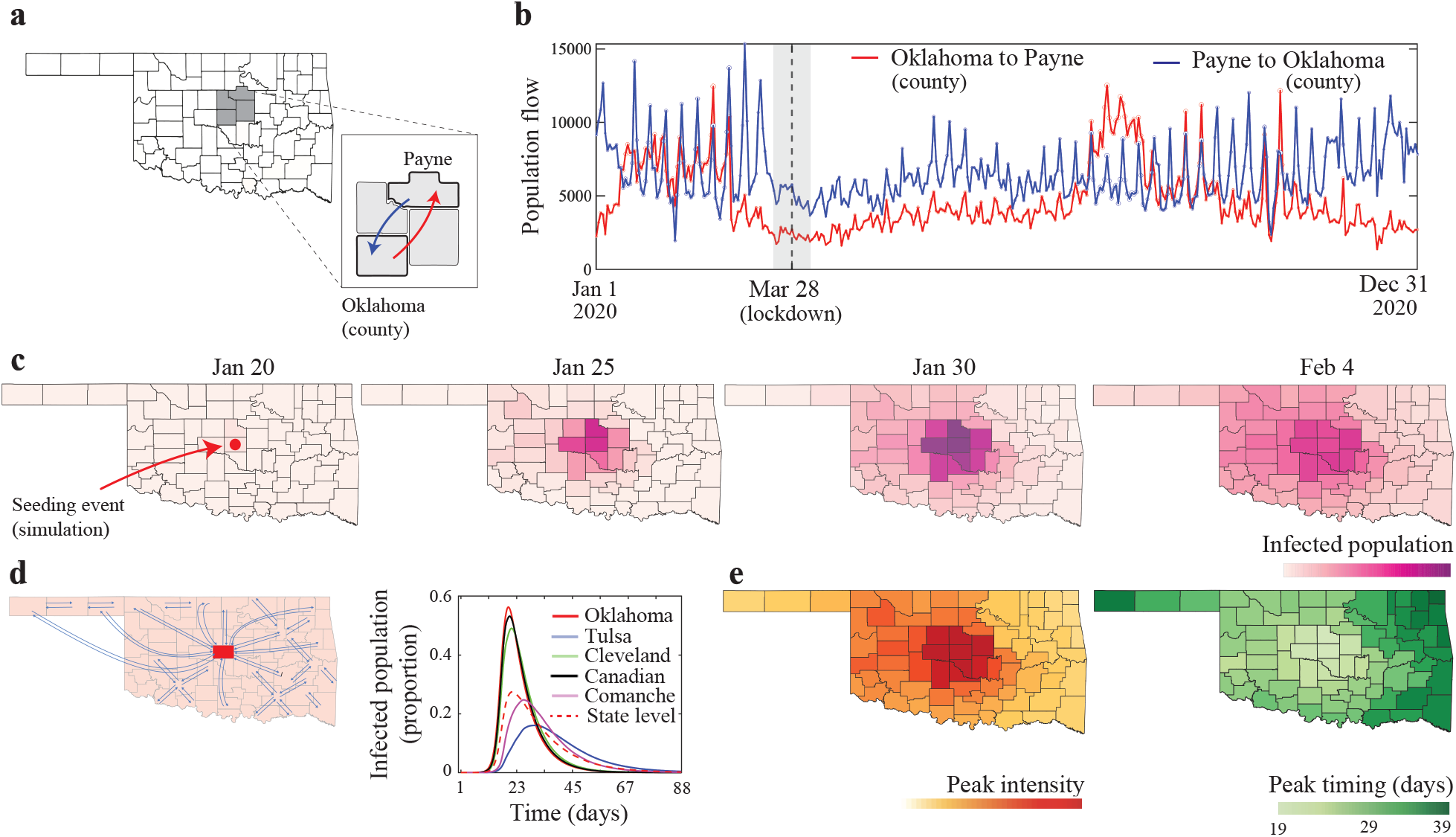
Integration of mobility data into an epidemic modeling framework. **a**, County boundaries for the state of Oklahoma. The inset illustrates Oklahoma and Payne counties. **b**, Daily population flows between Oklahoma and Payne counties from January to December 2020. **c**, Epidemic simulation seeded in Oklahoma County (red dot) and spatial-temporal dynamics at successive time points (January 20, January 25, January 30, and February 4 in 2020). **d**, Schematic network of inter-county mobility connections and normalized prevalence curves for representative counties compared to statewide aggregate (dashed line). **e**, County-level maps of peak epidemic intensity (maximum infected fraction) and peak timing (days since seeding). Parameter values: **c**, *β*_0_ = 0.1428 day^−1^ for all counties except the seeding county, where *β*_seed_ = 7 × *β*_0_. **d**, *β*_0_ = 0.20 day^−1^ for all counties except the seeding county, where *β*_seed_ = 7 × *β*_0_. See Table 1 for a summary of parameter values shared across all simulations.

To further explore the effects of mobility data, we compared the outputs of our model equipped with two alternative networks: a fully connected network with equal mobility levels, and a gravity model network [19] (see Methods and Extended Data Fig. 1). The fully connected structure produced highly synchronized and unrealistic epidemic curves across counties, failing to capture spatial variability in timing and intensity. In contrast, the gravity model introduced some degree of variability, but the resulting spatial patterns were still markedly smoother than those obtained with our mobility data.

**Table 1:** Key model parameters and initial conditions used in the simulations.

| Parameter | Description | Value / Source |
| --- | --- | --- |
| $M$ | Number of counties in the modeled state | State-specific (e.g., $M = 77$ for Oklahoma) |
| $\beta_j$ | Transmission rate for county $j$ | Variable; defined per scenario (see figure captions) |
| $\phi_{ij}(t)$ | Fraction of population in county $i$ traveling to county $j$ at time $t$ | Derived from mobility data (SafeGraph) |
| $N_i$ | Population size of county $i$ | SafeGraph-derived estimates |
| $\gamma$ | Recovery rate (inverse of infectious period) | Fixed at $0.1429 \text{ day}^{-1}$ (7-day infectious period) |
| $I_i(0)$ | Initial number of infectious individuals in county $i$ | $I_{\text{seed}}(0) = 3$; $I_i(0) = 0$ for all other counties |
| $S_i(0)$ | Initial number of susceptible individuals in county $i$ | $S_i(0) = N_i - I_i(0)$ for all counties |
| $R_i(0)$ | Initial number of recovered individuals in county $i$ | $R_i(0) = 0$ for all counties |

### Identifying epidemic hotspots to guide strategic control

Previous approaches have defined transmission or mobility hotspots based on density thresholds [25], population size and flows [15], or high-risk sites for disease importation [26]. Here, we introduce a novel algorithm that identifies epidemic hotspots as the counties most capable of driving statewide outbreaks, based on the combined effects of local transmission and network connectivity. More precisely, for a state with *M* counties, we simulated *M* epidemics, each seeded in a different county. The seeding county was assigned a five-fold higher transmission rate to hypothetically represent elevated contact activity and generate sufficiently large outbreaks within the pre-lockdown periods across states. To explore each county’s potential to trigger outbreaks under weak control, we set the baseline transmission rate in non-seeding counties slightly below the recovery rate (see Fig. 2 caption and Table 1 for parameter choices). We then computed the total epidemic size across the state resulting from each seeding scenario. Counties that generated disproportionately larger statewide outbreaks were identified as hotspots and colored by intensity (Fig. 2a, colors normalized per state). Our hotspot identification algorithm is described in detail in the Methods section (Algorithm 1).

**Figure 2:**
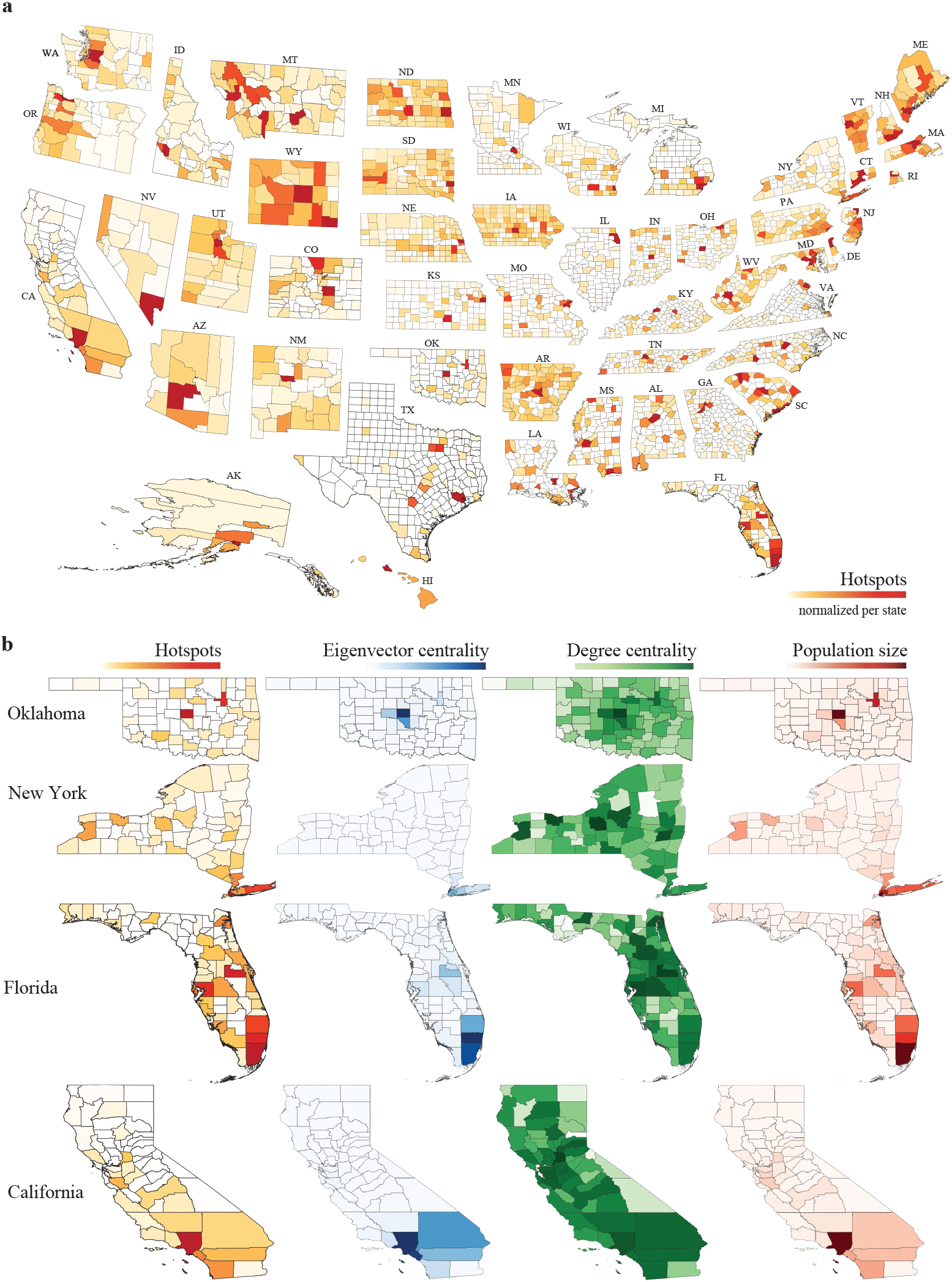
Epidemic hotspots in the United States. **a**, Hotspots identified using epidemic simulations (our approach). **b**, Comparison with network and population indicators in four representative states: Oklahoma, New York, Florida, and California. From left to right, maps show: hotspots identified via simulation (using the same metric as panel **a**); eigenvector and degree centrality of each county; and population size. Parameter values: **a**, *β*_0_ = 0.1428 day^−1^ for all counties except the seeding counties, where *β*_seed_ = 5 × *β*_0_. See Table 1 for a summary of parameter values shared across all simulations. **b**, See Methods for computation details on eigenvector and degree centralities.

We compared our hotspot definition to three standard indicators: (i) eigenvector centrality of the mobility network, (ii) degree centrality of the mobility network, and (iii) county population size (Fig. 2b). Eigenvector centrality and population size most closely aligned with our simulation-based hotspots, though our method yielded higher contrast, as reflected in a broader color range across counties in four representative states (Oklahoma, New York, Florida, and California). Degree centrality also identified many hotspot counties, though it tended to highlight a larger set of high-connectivity regions. These comparisons indicate that the most influential hotspots are both densely populated and well-connected. Such agreement among metrics reinforces the robustness of our approach, which offers a new method for identifying high-impact locations for epidemic spread. Supplementary Table S1 lists the top five hotspots alongside the top five counties ranked highest by each of the alternative metrics shown in Fig. 2b.

Our hotspot identification method takes into account the total epidemic size across the state, including infections within the seeding county. As a result, counties with large populations and intense local transmission can often be identified as hotspots. To evaluate each county’s ability to export infections beyond its borders, we further defined *superspreaders* by running simulations for each seeding county and computing the epidemic size across the state, excluding the seeding county itself (see Methods, Algorithm 1). This metric quantifies a county’s role as a hub for outward transmission. We found a strong agreement between superspreaders and hotspots. In most states, the two sets were nearly identical (see Arkansas, Extended Data Fig. 2a, and Supplementary Tables S2, S3, and S4). In a few exceptions—such as Missouri—counties like Gentry and Shelby emerged as superspreaders despite not being identified as hotspots (Extended Data Fig. 2b). These findings highlight that hotspots tend to be densely populated and retain the capacity to drive widespread transmission beyond their borders.

### Rethinking lockdowns through targeted interventions

Having identified epidemic hotspots, we utilized our mobility-informed metapopulation model to simulate targeted intervention strategies. Since hotspots can amplify statewide transmission, we tested two classes of targeted interventions: (i) locally reducing the baseline basic reproduction number (*R*_0_) at hotspot counties by a scaling factor *η*, (Fig. 3a), and (ii) restricting mobility by scaling outbound flow from hotspots by a factor *α* (Fig. 3b). The parameters *η* and *α* map directly to public health actions: reductions in local contact rates and controls on travel from hotspots.

**Figure 3:**
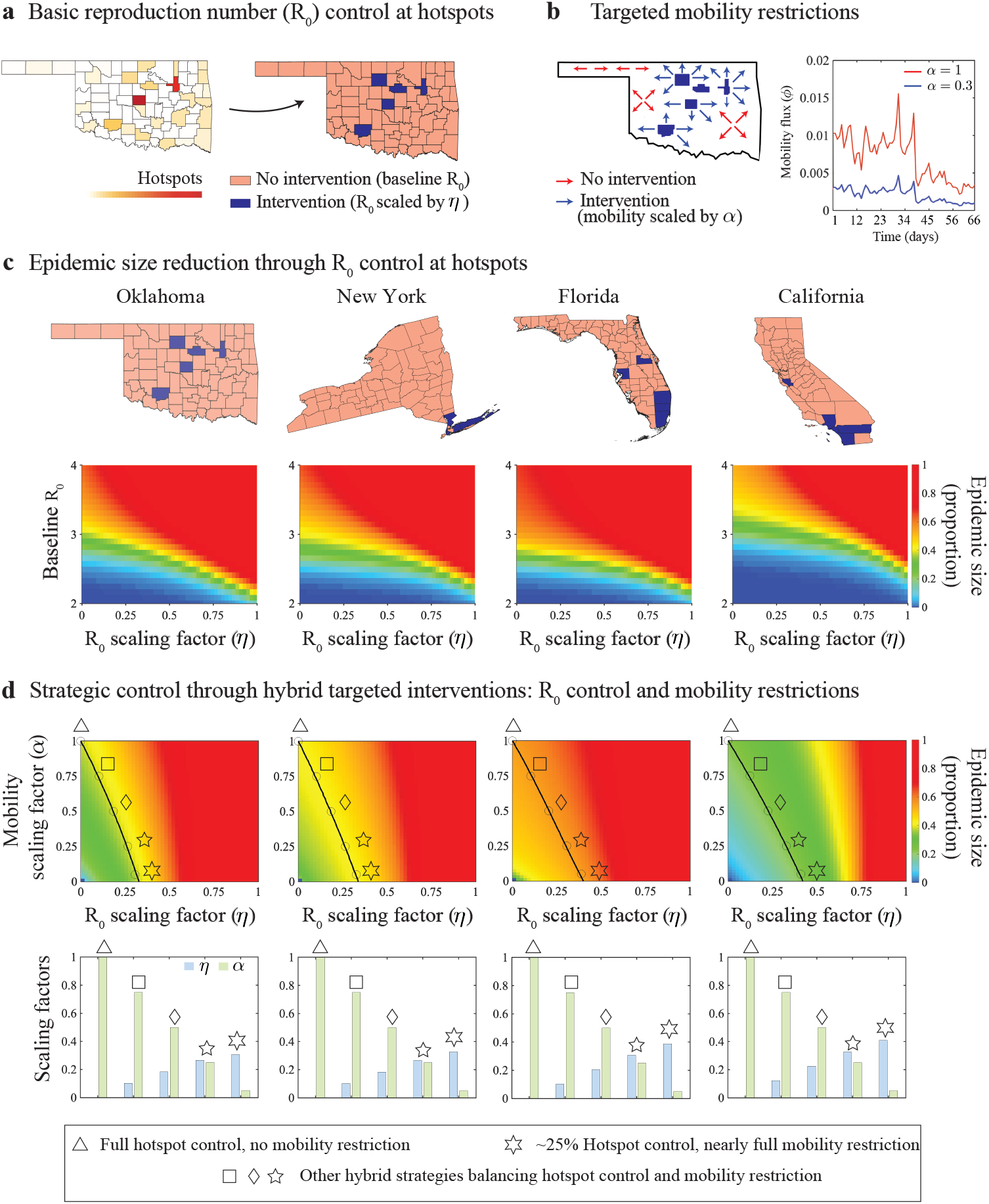
Targeted interventions as an alternative to blanket epidemic control. **a**, Localized interventions reduce the basic reproduction number (*R*_0_) at the top five hotspot counties (dark blue), compared to surrounding counties with baseline *R*_0_ (pink), by scaling transmission with factor *η*. **b**, Mobility-based interventions reduce mobility from hotspots to other counties by scaling with factor *α*. Right: time series of mobility flux (*ϕ*) under no restriction (*α* = 1) and partial restriction (*α* = 0.3). **c**, Epidemic size as a function of baseline *R*_0_ and hotspot scaling factor *η* for four representative states (Oklahoma, New York, Florida, California). Color gradients show the total statewide epidemic size, from uncontrolled spread (red) to near-complete suppression (∼90% reduction, blue). **d**, Hybrid strategies combine hotspot *R*_0_ control and mobility restrictions when baseline *R*_0_ = 3. Colormaps display epidemic size as a function of *η* and *α*; black contours denote equivalent epidemic outcomes. Symbols mark representative strategies: full hotspot suppression with no mobility restriction (triangle), nearly full mobility restriction with partial hotspot control (six-point star), and balanced hybrid strategies (diamond, square, five-point star) achieving equivalent control. Bar plots show the trade-offs between *η* and *α* for each strategy. Seeding counties for panel **c**: Oklahoma County (OK), New York County (NY), Miami-Dade (FL), Los Angeles County (CA). See Table 1 for parameter values.

We first evaluated interventions that reduced hotspot *R*_0_ values across a range of transmission scenarios in four representative states: Oklahoma, New York, Florida, and California (Fig. 3c). To simulate these interventions, we ran simulations in which epidemics were seeded in the biggest hotspot for a range of baseline *R*_0_ values. Specifically, we set a baseline transmission rate *β*_0_ at all non-hotspot counties, while assigning each hotspot a scaled transmission rate *η × β*_0_, keeping the recovery rate *γ* fixed (see Table 1). Colormaps in Fig. 3c depict the proportion of the statewide population infected under varying baseline *R*_0_ and scaling factors *η*, from complete suppression of local transmission (*η* = 0) to no intervention (*η* = 1). Targeted interventions at hotspots significantly reduced statewide epidemics, indicating that blanket statewide interventions may not be required. In the colormaps, yellow regions correspond to a normalized epidemic size of ∼0.4, representing a 60% reduction compared to uncontrolled spread, while blue regions indicate near-complete suppression, with epidemic sizes close to zero (∼90% reduction). Consistent patterns across four diverse states confirm the robustness of our framework, with some regional variability. For instance, California exhibited broader regions of epidemic suppression compared to Florida, revealing regional differences in the efficacy of interventions. Importantly, controlling hotspots was critical across seeding scenarios. By contrast, interventions targeting *coldspot* counties consistently failed, resulting in persistent large outbreaks for different scaling factors and seeding counties (See Methods and Extended Data Fig. 3).

Next, we simulated hybrid interventions that simultaneously reduce both *R*_0_ and outbound mobility at hotspot counties under a baseline transmission scenario of *R*_0_ = 3, representing a highly transmissible respiratory disease such as COVID-19 early in the pandemic (Fig. 3d). Colormaps display the proportion of the statewide epidemic size as a function of the scaling factors *η* and *α*, each ranging from 0 (complete control) to 1 (no intervention). Here, *α* = 0 corresponds to a complete elimination of mobility from hotspots. Overlaid black contour lines denote combinations of *η* and *α* that yield the same epidemic outcomes, which vary by state. We highlight five representative strategies using distinct symbols. The triangle indicates a scenario where hotspot transmission is fully suppressed (*η* = 0), while mobility remains unrestricted (*α* = 1). This reflects a strict and idealized local control effort. Notably, several hybrid strategies—marked by square, diamond, and five-point star—achieve the same level of epidemic control with moderate local *R*_0_ suppression (0.1 *< η <* 0.3, corresponding to a 10–30% reduction) combined with partial mobility restrictions (0.3 *< α <* 0.8, corresponding to a 30–80% reduction). These hybrid strategies reveal practical trade-offs that reduce the societal burden of overly stringent single-measure interventions. The six-point star denotes a complementary strategy where mobility from hotspots is almost fully restricted, while hotspot transmission is even less controlled (*η* ∼ 0.25 in Oklahoma and New York, *η* ranging from 0.25 to 0.5 in Florida and California).

## Discussion

Our findings demonstrate that strategic county-level interventions can achieve statewide epidemic control, offering an alternative to blanket responses. As the world continues to confront emerging pathogens [27, 28], effective preparedness requires designing interventions that deliver maximum impact while reducing social and economic burden.

Our mobility-informed metapopulation model integrates comprehensive data from GPS-enabled mobile devices to parametrize realistic movement networks. Numerical simulations revealed geographic patterns of disease spread, demonstrating our model’s ability to reproduce transmission dynamics influenced by human mobility. Using this computational framework, we identified epidemic hotspots as counties with the greatest potential to amplify transmission and drive statewide outbreaks. We then assessed whether strategic, localized interventions could match the effectiveness of broad, aggressive measures. Our results demonstrate that targeted *R*_0_ reductions at hotspot counties alone can significantly curb statewide outbreaks. Moreover, hybrid strategies – moderately reducing both local *R*_0_ and outbound mobility at hotspots – can replicate the effects of full local suppression (*R*_0_ = 0 at hotspots), offering a more adaptable and less burdensome path to epidemic control.

We built upon previous work [13, 29, 30] by extending the classical SIR framework [20] into a transparent and interpretable metapopulation model parameterized with empirically derived population flows, rather than theoretical mobility structures such as gravity models [19]. This design preserves mathematical transparency and interpretability, offering insight into how human movement shapes epidemic dynamics. By avoiding the complexity of heavily parameterized models, our framework maintains a direct mapping between model structure and policy levers—specifically, local transmission rates and mobility restrictions. Our hotspot identification algorithm provides a principled assessment that complements traditional indicators such as centrality metrics and population size. While eigenvector centrality and population size most closely aligned with our identified hotspots, our approach yielded a sharper contrast, reflecting the interplay between transmission and network connectivity that static measures fail to capture.

In contrast to recent studies [8, 14, 16], we avoided numerical calibration to specific epidemic curves, instead emphasizing general principles and qualitative insights into disease dynamics. Future work could investigate our model’s potential for data fitting, which would require pathogen-specific parameters such as latency and waning immunity periods. While in this study, we focused on interventions from an epidemiological perspective, decision-makers may also weigh the economic value of particular regions. Coupling mobility-informed transmission models with regional economic data could help identify trade-offs between public health goals and economic stability [31]. Realizing such a framework represents a promising direction for expanding the scope and utility of mobility-based epidemic models.

Our approach has limitations. The model does not account for behavioral feedback on local transmission rates, which are held constant over time. While this assumption may be reasonable during the early stages of an outbreak, it likely becomes unrealistic once public health interventions and behavioral responses begin to influence transmission dynamics. The underlying network model assumes instantaneous return travel, capturing average daily flow dynamics but not the temporal structure of movement represented in simple-trip models [3, 32]. This simplification likely increases effective mixing between populations compared to real-world mobility, where travel and return occur on longer time scales. Nevertheless, the fact that our targeted interventions reduce statewide outbreak sizes even under these heightened mixing conditions suggests that the qualitative patterns we observe are robust. Each state is treated as an isolated system, excluding interstate and air travel. Incorporating importation risk—particularly into counties with airports—could improve realism by accelerating transmission in key travel hubs during early outbreak phases. The mobility data used in our study also carries known limitations, including incomplete population coverage, sampling biases, and the lack of cross-state flow information [33, 34, 35, 36, 37]. Yet these data have proven valuable in epidemic modeling, offering temporally resolved and behaviorally grounded movement patterns [26, 37, 38, 39]. In contexts where such empirical data are unavailable, gravity models remain a viable alternative. We selected the county level as our spatial scale, reflecting the level at which many public health decisions are implemented in the United States. Finer resolutions, such as census block groups, may reveal more granular patterns [40]. Future studies could explore a similar framework at other geographic scales.

Our framework provides a transparent and interpretable tool for pandemic preparedness, addressing the pressing need for actionable models to evaluate localized control strategies. As public health systems struggle with growing complexity and uncertainty in pandemic preparedness, our modeling tool strikes a balance between simplicity, realism, and policy relevance. Our approach allows forward-looking evaluation of intervention strategies and demonstrates that statewide control can be achieved without resorting to overly restrictive measures.

## Data Availability

The human mobility data used in this study are publicly available on GitHub at
https://github.com/GeoDS/COVID19USFlows. The simulation codes that support the find-
ings of this study are available upon reasonable request.

## Methods

### Mobility data and flux matrices

The human mobility data was obtained from SafeGraph, which tracks approximately 10% of all GPS-enabled mobile devices in the United States. Dynamic origin-to-destination (OD) population flows were generated by analyzing the visit patterns of mobile device users to various points of interest [22, 23]. SafeGraph data has been validated against alternative mobility sources, such as Google’s mobility reports [14], and has been extensively used in recent epidemiological studies, particularly during the COVID-19 pandemic [41, 42, 43].

We model each U.S. state as an isolated metapopulation, where counties serve as nodes representing both origins and destinations of human movement. To keep the model tractable and better identify hotspot counties for each state, we assume that daily movement between counties in different states is negligible. The population flow from county *O*_*i*_ (origin, indexed by *i*) to county *D*_*j*_ (destination, indexed by *j*) on day *t* is denoted as *F* [*i*][*j*][*t*]. Fig. 1b exhibits a representative time series of population flows from Oklahoma County (origin) to Payne County (destination) in the state of Oklahoma during the year 2020. Throughout this study, we simulated epidemic dynamics using mobility data from January 1, 2020, up to the day before each state implemented its first COVID-19 or stay-at-home order (Tables S5 and S6). This temporal cutoff ensured that all simulations reflect pre-pandemic mobility patterns before policy interventions began to alter movement. As such, our goal is not to reproduce the COVID-19 pandemic *per se*, but to explore epidemic scenarios under typical, unperturbed mobility conditions as a foundation for general pandemic preparedness.

For each pair of counties *i* and *j*, the flux matrix entry *ϕ*_*ij*_(*t*) represents the fraction of the population in county *i* that traveled to county *j* on day *t*, and is constrained to the interval [0, 1]. For *i* ≠ *j*, we define 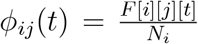, where *N*_*i*_ is the total population of county *i*. The diagonal element *ϕ*_*ii*_(*t*), denoting the fraction of the population remaining in the origin county, is computed as *ϕ*_*ii*_(*t*) = 1− ∑ _*j*≠*i*_ *ϕ*_*ij*_(*t*), provided that ∑_*j*≠*i*_ *F* [*i*][*j*][*t*] ≤ *N*_*i*_. In the rare cases where ∑_*j*≠*i*_ *F* [*i*][*j*][*t*] *> N*_*i*_—a result of multiple same-day trips by individual users—we use the total outgoing flow ∑ _*j≠i*_ *F* [*i*][*j*][*t*] as the normalization factor and set *ϕ*_*ii*_(*t*) = 0 for simplicity. By calculating each entry *ϕ*_*ij*_(*t*), we constructed time-dependent flux matrices

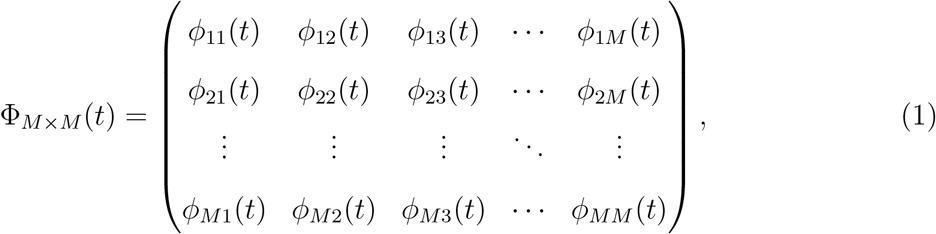

which were directly integrated into our model to simulate mobility-driven epidemic dynamics across counties.

### Flux matrix for a fully connected network

We utilized the flux matrix structure (equation 1) to define mobility networks with varying connectivity patterns. One comparison involved simulations based on Oklahoma data versus a fully connected network, in which all counties are assumed to be equally connected (Extended Data Fig. 1). In this configuration, all off-diagonal fluxes are constant and set to a value *ϕ*_0_ ∈ (0, 1), while diagonal entries are adjusted to ensure row normalization:

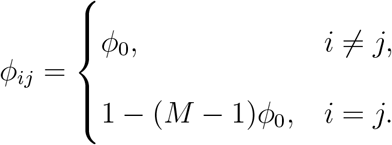

For these simulations, we used *ϕ*_0_ = 0.0068, corresponding to the average off-diagonal entry of the flux matrix during the pre-lockdown period in the state of Oklahoma.

### Flux matrix from gravity models

We also compared our mobility-informed model with a classic gravity model [19] (Extended Data Fig. 1). The gravity model estimates the average number of travelers between two locations as:

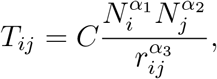

where *C* is a proportionality constant, *α* and *β* control the influence of origin and destination population sizes, and *γ* governs the distance-based decay of mobility flows. These parameters vary by region and are estimated through calibration.

The *T*_*ij*_ values serve as synthetic analogs to the empirical mobility matrix *F* [*i*][*j*][*t*] derived from mobility data. To compute the gravity-based flux matrix Φ^*GM*^, we normalized *T*_*ij*_ by dividing by max 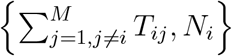, yielding off-diagonal fluxes 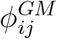. Diagonal elements were calculated to ensure row-normalization: 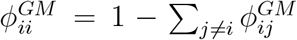. We calibrated the parameters *C, α*_1_, *α*_2_, *α*_3_ such that the resulting flux matrix entries approximate the time averaged empirical flux matrix ⟨Φ(*t*)⟩_*t*_, by minimizing the Frobenius norm of their difference: ∥Φ^*GM*^ − ⟨Φ(*t*)⟩_*t*_∥_*F*_ [44]. We initialized the optimization with parameters from Engebretsen et al. [45]: [*C, α*_1_, *α*_2_, *α*_3_] = [0.404, 0.690, 0.688, 0.946]. The final calibrated values were *C* = 0.00166, *α*_1_ = 0.74149, *α*_2_ = 0.84953, and *α*_3_ = 1.

### Mobility-informed metapopulation model

We modeled disease transmission dynamics using a mobility-informed SIR network model [29, 30], in which each U.S. state is treated as an isolated network of *M* counties connected by mobility fluxes. Our model describes the temporal dynamics of susceptible, infected, and removed populations in each county through the following system of differential equations:

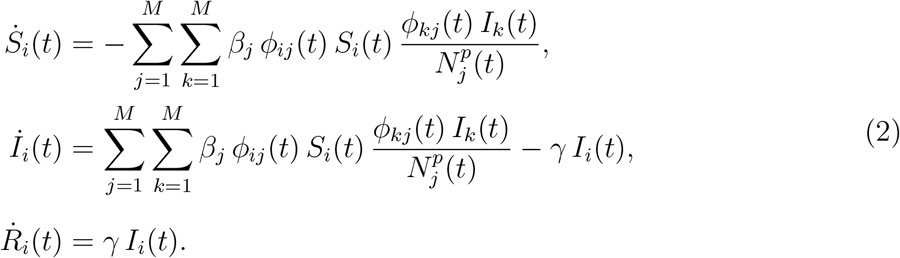

Here *S*_*i*_(*t*), *I*_*i*_(*t*), and *R*_*i*_(*t*) denote the number of susceptible, infected, and removed individuals in county *i* at time *t*, respectively. The total population of county *i* is denoted *N*_*i*_, with a county-specific transmission rate *β*_*i*_ and a recovery rate *γ*. In our simulations, we generally assumed uniform transmission rates across all counties except for the seeding county. Specifically, we set *β*_*j*_ = *β*_0_ for all counties *j* except the seeding county, where we defined *β*_seed_ = *ζβ*_0_ for a scaling factor *ζ >* 1. For hotspot identification, we chose *ζ* = 5 to represent a consistently higher transmission rate due to elevated contact activity. In simulations designed to highlight the influence of mobility data in our model, we used *ζ* = 7 to further amplify spatial propagation patterns during the early stages of the epidemic. The *present* population in county *j* at time *t* is a dynamic variable, accounting for incoming mobility, and is given by 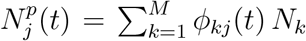. Table 1 presents the parameter values, their descriptions, and the corresponding data sources used in our simulations.

### Hotspot identification

To identify epidemic hotspots within each U.S. state, we systematically simulated our mobility-informed metapopulation model with initial seeding events in every county. In each simulation, the seeding county was assigned a five-fold higher transmission rate to emulate elevated contact activity and produce outbreaks of sufficient magnitude within the pre-lockdown windows. We then computed the total statewide epidemic size resulting from each seeding event. To identify superspreaders, we performed a similar analysis, excluding infections that occurred within the seeding county itself. Algorithm 1 outlines the procedure for determining the top five hotspots and superspreaders. To find coldspots, we simply take the last five locations in the Normalized epidemic size vector 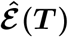 (Extended Data Fig. 3). We compared the obtained hotspots with eigenvector and degree centrality using the same mobility network employed in the model (see Fig. 2b and Supplementary Table S1).

### Centrality Measures of the Mobility Network

We compared the hotspots obtained from epidemic simulations with two standard network metrics derived from cellphone mobility data: (i) degree centrality (DC) and (ii) eigenvector centrality (EC). In directed networks, the degree of a node can be decomposed into in-degree (incoming connections) and out-degree (outgoing connections). For each day *t*, we calculated the out-degree centrality of county *i* as the number of counties *j* such that *F* [*i*][*j*][*t*] *>* 0, divided by *M* − 1, where *M* is the number of counties in the state. Similarly, in-degree centrality was computed by counting the number of counties *j* with *F* [*j*][*i*][*t*] *>* 0, also normalized by *M* − 1. We then averaged the in-degree and out-degree centralities over the pre-lockdown period and computed the final degree centrality for county *i* as

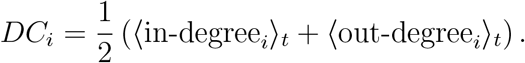

#### Algorithm 1

Top five hotspot/superspreader identification

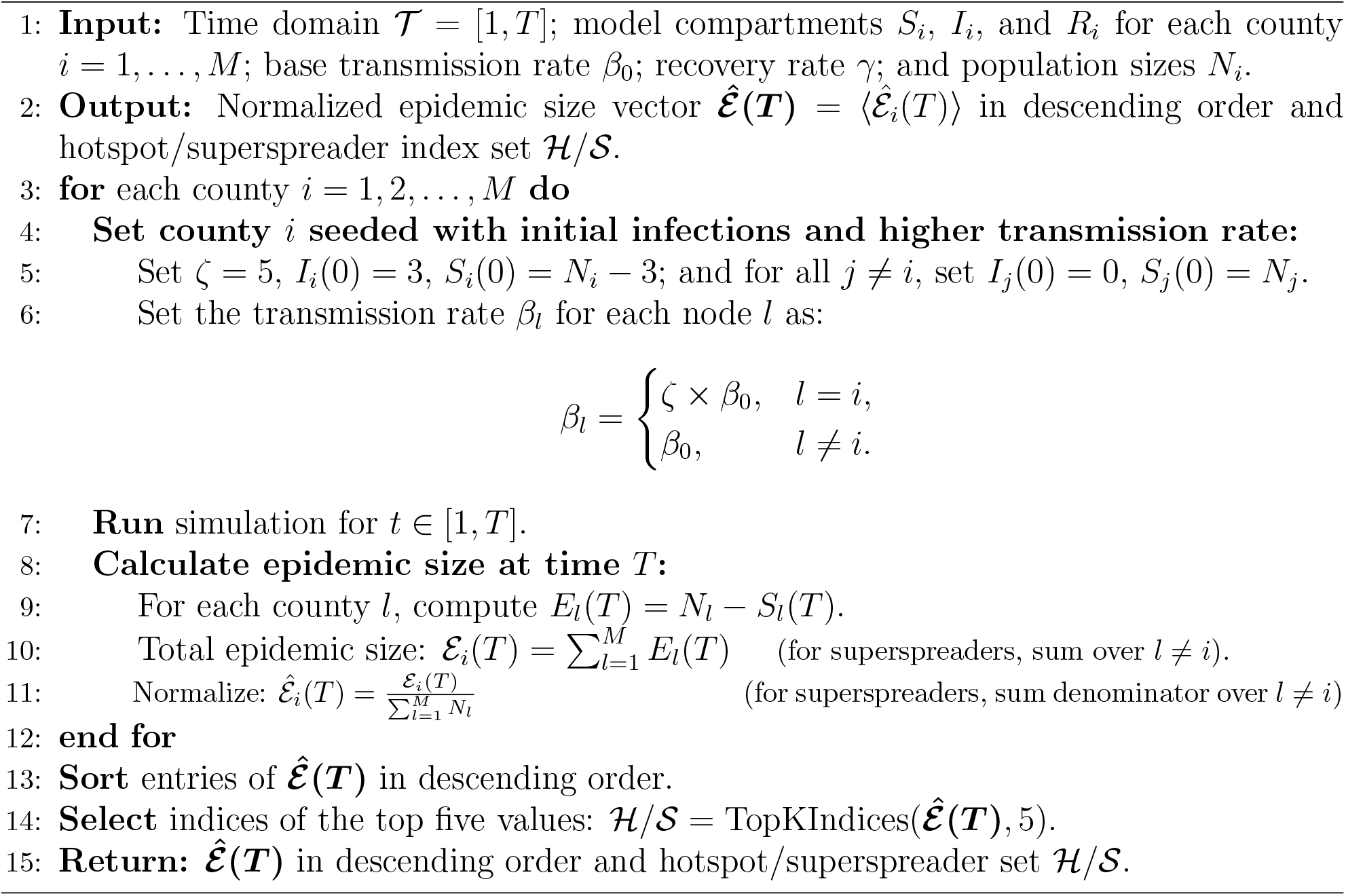

Eigenvector centrality assigns higher importance to nodes that are connected to other highly connected nodes [46, 47]. This property makes it particularly well-suited for identifying counties that are not only well-connected but also strategically positioned within the broader network of influential nodes, regardless of whether the network is directed or undirected. Mathematically, let *A* = (*a*_*ij*_) denote the adjacency matrix of a graph. The eigenvector centrality *x*_*i*_ of node *i* is defined as:

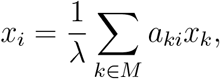

where *λ* ≠ 0 is a constant. In matrix form, this relationship becomes *λ***x** = *A***x**, which implies that the centrality vector **x** is a right-hand eigenvector of the adjacency matrix *A*, corresponding to eigenvalue *λ*. The eigenvector associated with the largest eigenvalue is used to compute centrality scores. Using the `NetworkX` library [48, 49], we computed daily eigenvector centrality values from the mobility network defined by the population flow matrices *F* [*i*][*j*][*t*] during the pre-lockdown period. The final centrality score for each county was computed as the temporal average, i.e., *EC*_*i*_ = ⟨*x*_*i,t*_⟩_*t*_.

## Data and Code Availability

The human mobility data used in this study are publicly available on GitHub at https://github.com/GeoDS/COVID19USFlows. The simulation codes that support the findings of this study are available upon reasonable request.

## Acknowledgments

L.M.S. and T.H. acknowledge support from the National Science Foundation (NSF DMS-2327844). M.S. was partially funded by the cooperative agreement CDC-RFA-FT-23-0069 from the CDC’s Center for Forecasting and Outbreak Analytics.

## Author contributions

H.K.D. processed the mobility data, designed numerical experiments, performed simulations, analyzed results, generated figure panels, designed numerical experiments, and drafted portions of the Methods section. T.H. curated the mobility data and contributed to drafting the Methods section. M.S. provided conceptual guidance, critical feedback on the modeling framework, and contributed to writing and revising the manuscript. L.M.S. conceived and supervised the project and drafted the initial version of the manuscript. All authors reviewed, edited, and approved the final manuscript.

**Extended Data Fig1:**
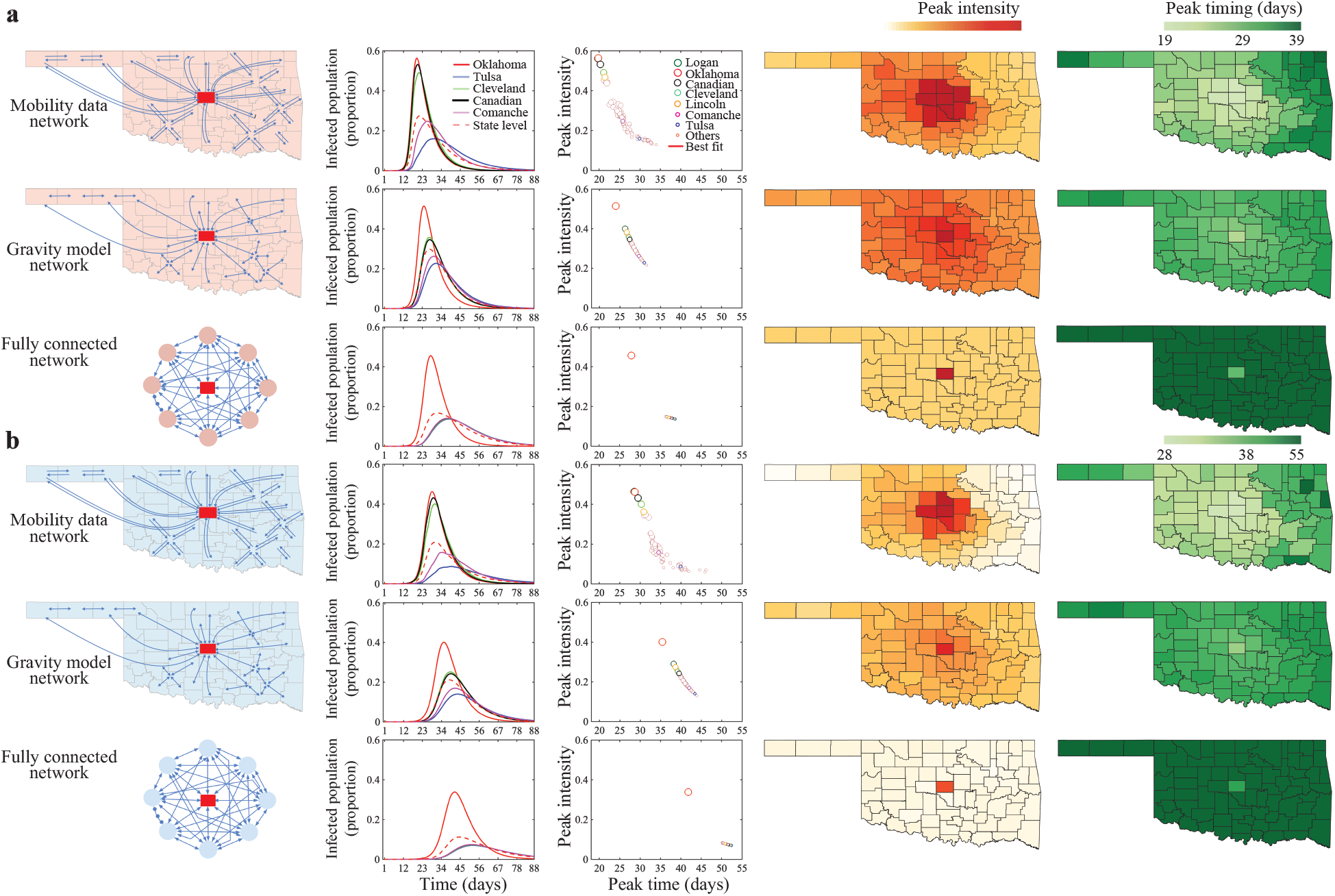
Comparison of simulated epidemics under different mobility networks. We assessed epidemic dynamics in Oklahoma using our mobility-informed metapopulation model, and compared results under three network structures: (i) our mobility networks from GPS-enabled mobile devices, (ii) a gravity model, and (iii) a fully connected network with uniform mobility (see Methods). Epidemics were seeded in Oklahoma County, which was assigned a higher transmission rate relative to other counties. **a**, Simulation results when the baseline transmission rate *β*_0_ exceeds the recovery rate (*β*_0_ *> γ*). **b**, Results when *β*_0_ *< γ*. In both cases, the mobility-informed model exhibits greater spatial variability in epidemic peak timing and intensity compared to gravity-based and fully connected networks. For example, the peak timing vs. intensity plots (third column) show a marked reduction in spatial variability under gravity and fully connected networks compared to the mobility-informed network, where points are more widely dispersed. For the fully connected network, we used a uniform flux *ϕ*_0_ = 0.0068, matching the average pre-lockdown mobility flux observed in Oklahoma during 2020. Parameter values **a**, *β*_0_ = 0.20 day^−1^ for all counties except the seeding county, where *β*_seed_ = 7 *× β*_0_. **b**, *β*_0_ = 0.1428 day^−1^ for all counties except the seeding county, where *β*_seed_ = 7 × *β*_0_. See Table 1 for a summary of parameter values shared across all simulations. See Methods for details on Fully Connected and Gravity model networks.

**Extended Data Fig2:**
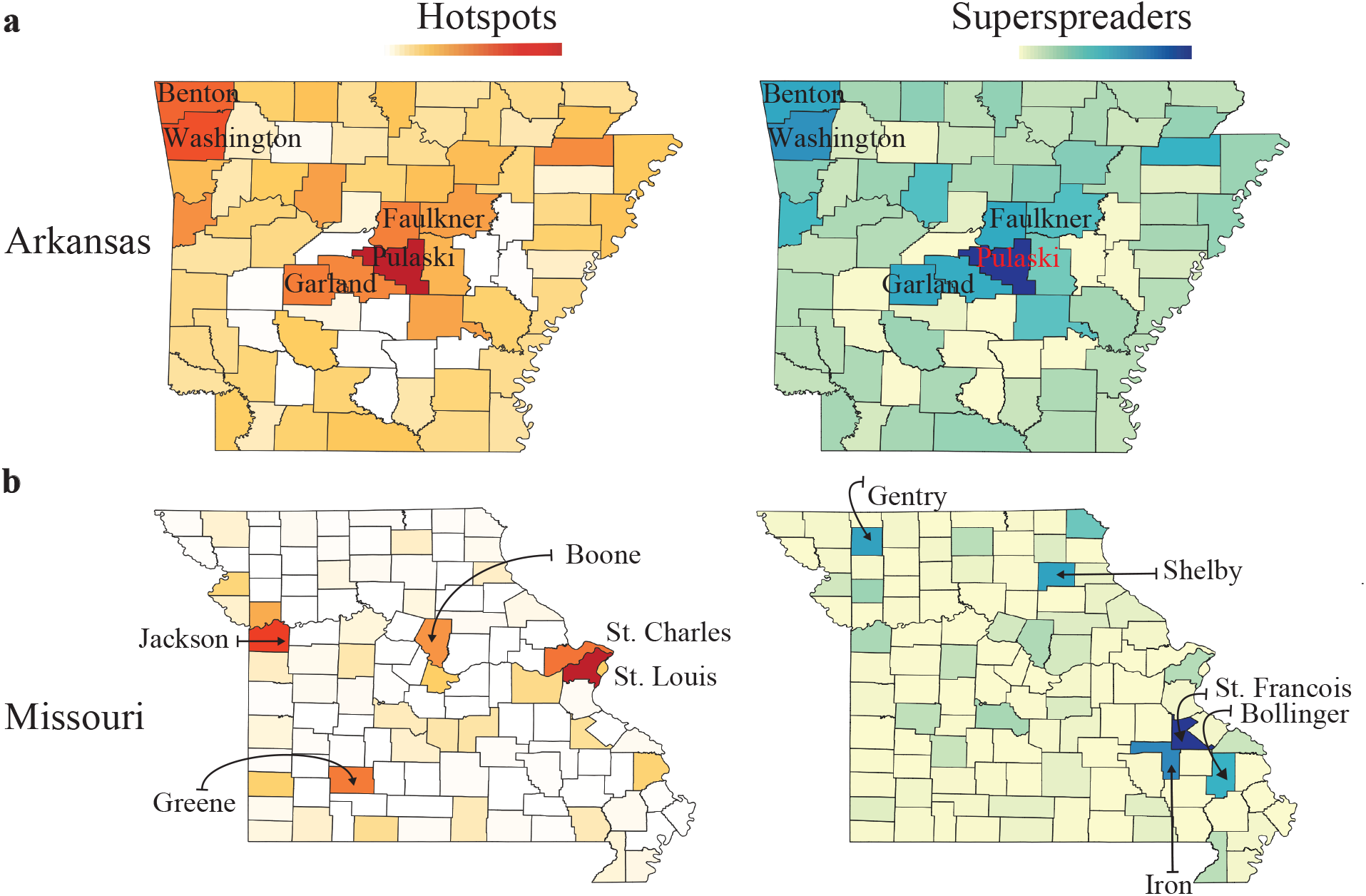
Comparison of epidemic hotspots and superspreaders: We compared two complementary metrics of epidemic influence across counties: hotspots, defined by the total statewide epidemic size when a county is seeded with elevated transmission; and superspreaders, defined by the epidemic size in the rest of the state, excluding the seeding county (see Methods). **a**, In Arkansas, the two measures largely agree, with populous and central counties like Pulaski and Garland identified under both metrics. **b**, In Missouri, discrepancies emerge—counties such as Gentry and Shelby are classified as superspreaders despite not being hotspots. In most states, the two sets were nearly identical (Supplementary Tables S2, S3, and S4). Parameter values: **a**,**b** *β*_0_ = 0.1428 day^−1^ for all counties except the seeding counties, where *β*_seed_ = 5 *× β*_0_. See Table 1 for a summary of parameter values shared across all simulations.

**Extended Data Fig3:**
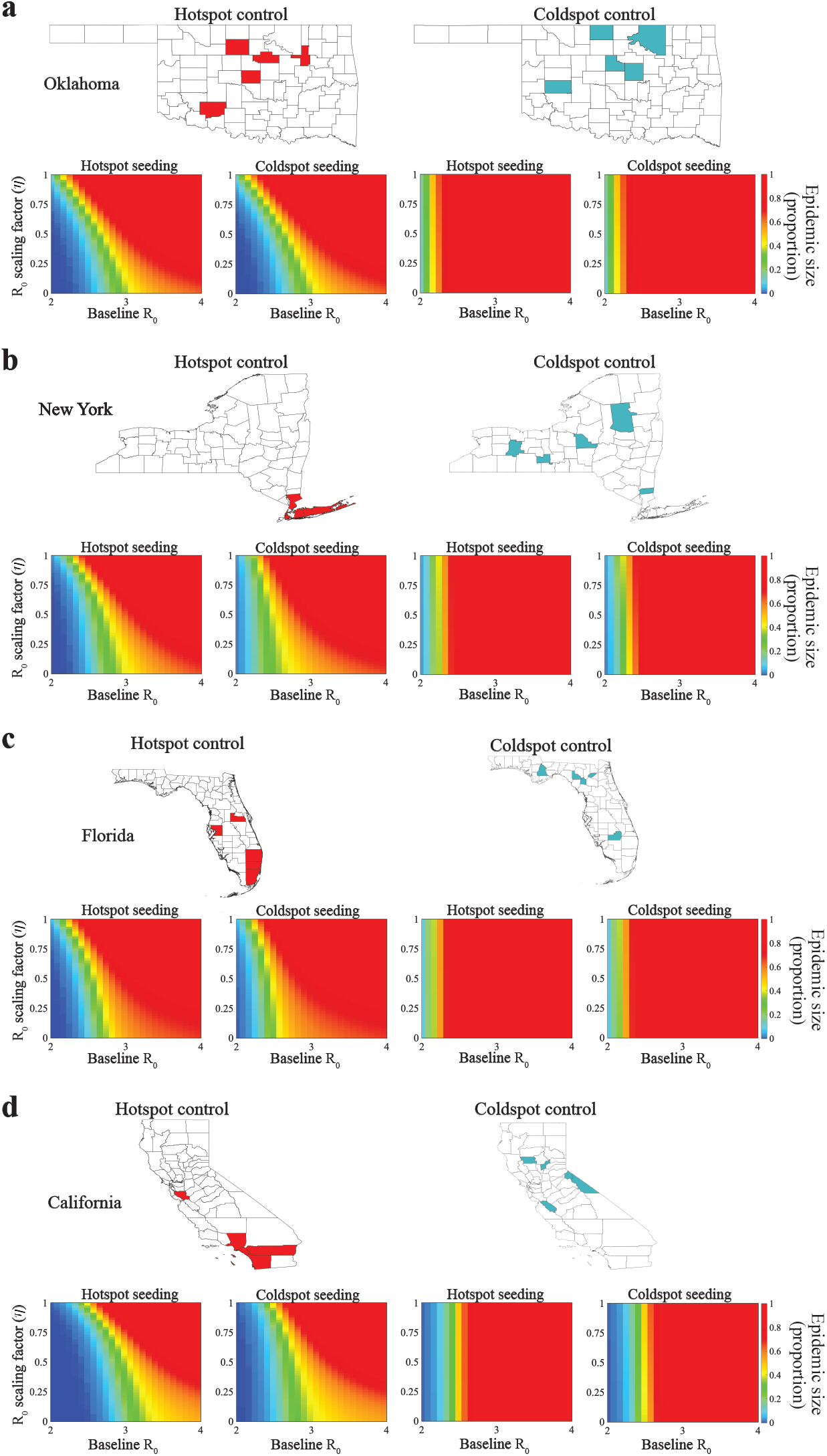
Effectiveness of targeted interventions at hotspots versus coldspots. To evaluate the importance of hotspot targeting, we compared intervention outcomes when control efforts were applied either to identified hotspots or to randomly selected coldspots. Epidemics were seeded in both hotspot and coldspot counties, and outcomes were simulated across a range of baseline transmission rates (*R*_0_) and hotspot control intensities (scaling factor *η*; see Methods). **a–d**, Panels for Oklahoma, New York, Florida, California. Subpanels display epidemic sizes as a function of *R*_0_ and *η* under four scenarios: seeding in and controlling hotspots, seeding in and controlling coldspots, and their cross-combinations. Hotspots are shown in red and coldspots in cyan. Seeding counties: **a**, Oklahoma County (hotspot) and Logan (coldspot); **b**, New York County (hotspot) and Hamilton (coldspot); **c**, Miami-Dade (hotspot) and Glades (coldspot); **d**, Los Angeles County (hotspot) and Alpine (coldspot). See Table 1 for parameter values.

## Supplementary Information

**Table S1:** For four representative states (Oklahoma, New York, Florida, and California), we report the top five counties identified as epidemic hotspots, alongside the top five counties ranked by eigenvector centrality, degree centrality, and population size.

| State | Hotspots | Eigenvector Centrality | Degree Centrality | Population Size |
| --- | --- | --- | --- | --- |
| OK | Oklahoma, Tulsa, Comanche, Payne, Garfield | Oklahoma, Cleveland, Canadian, Tulsa, McClain | Oklahoma, Cleveland, Tulsa, Canadian, Payne | Oklahoma, Tulsa, Cleveland, Canadian, Comanche |
| NY | New York, Kings, Suffolk, Westchester, Nassau County | New York, Queens, Kings, Bronx, Nassau | Onondaga, Monroe, Albany, Erie, Oneida | Kings, Queens, New York, Suffolk, Bronx |
| FL | Miami-Dade, Orange, Broward, Hillsborough, Palm Beach County | Broward, Miami-Dade, Palm Beach, Orange, Seminole County | Orange, Hillsborough, Marion, Duval, Polk County | Miami-Dade, Broward, Hillsborough, Palm Beach, Orange |
| CA | Los Angeles, Orange, San Diego, Santa Clara, Riverside | Los Angeles, Orange, San Bernardino, Riverside, San Diego | Sacramento, Los Angeles, Alameda, Placer, Santa Clara | Los Angeles, San Diego, Orange, Riverside, San Bernardino |

**Table S2:** Top five hotspots and superspreaders across the United States (AL-GA).

| **FIPS** | **State** | **Hotspots** | **Superspreaders** |
| --- | --- | --- | --- |
| 01 | AL | Jefferson, Madison, Montgomery, Mobile, Tuscaloosa | Jefferson, Madison, Montgomery, Tuscaloosa, Mobile |
| 02 | AK | Anchorage, Matanuska-Susitna, Fairbanks North Star, Kenai Peninsula, Juneau City and Borough | same as hotspots |
| 04 | AZ | Maricopa, Pima, Yavapai, Coconino, Mohave | same as hotspots |
| 05 | AR | Pulaski, Washington, Benton, Garland, Faulkner | Pulaski, Washington, Garland, Benton, Faulkner |
| 06 | CA | Los Angeles, Orange, San Diego, Santa Clara, Riverside | same as hotspots |
| 08 | CO | El Paso, Larimer, Boulder, Denver, Pueblo | same as hotspots |
| 09 | CT | New Haven, Hartford, Fairfield, New London, Windham | same as hotspots |
| 10 | DE | New Castle, Kent, Sussex | same as hotspots |
| 12 | FL | Miami-Dade, Orange, Broward, Hillsborough, Palm Beach | Orange, Miami-Dade, Hillsborough, Broward, Palm Beach |
| 13 | GA | Fulton, Gwinnett, Cobb, Chatham, Muscogee | Fulton, Gwinnett, Chatham, Cobb, Muscogee |

**Table S3:** State-wise top five hotspots and top five superspreaders across the United States (HI-NJ).

| FIPS | State | Hotspots | Superspreaders |
| --- | --- | --- | --- |
| 15 | HI | Honolulu, Hawaii, Maui, Kauai, Kalawao | same as hotspots |
| 16 | ID | Ada, Canyon, Bonneville, Bannock, Twin Falls | Ada, Canyon, Bonneville, Twin Falls, Bannock |
| 17 | IL | Cook, Lake, Champaign, Winnebago, Sangamon | same as hotspots |
| 18 | IN | Marion, Allen, Lake, Tippecanoe, St. Joseph | Marion, Allen, Tippecanoe, Lake, Monroe |
| 19 | IA | Polk, Linn, Johnson, Black Hawk, Story | same as hotspots |
| 20 | KS | Johnson, Sedgwick, Shawnee, Douglas, Wyandotte | same as hotspots |
| 21 | KY | Jefferson, Fayette, Warren, Hardin, Boone | same as hotspots |
| 22 | LA | East Baton Rouge, Orleans, Jefferson, Lafayette, St. Tammany | East Baton Rouge, Orleans, Lafayette, Jefferson, St. Tammany |
| 23 | ME | Cumberland, Penobscot, York, Kennebec, Androscoggin | Cumberland, Penobscot, Kennebec, York, Androscoggin |
| 24 | MD | Prince George's, Montgomery, Baltimore, Anne Arundel, Baltimore city | Prince George's, Montgomery, Anne Arundel, Baltimore, Baltimore city |
| 25 | MA | Middlesex, Worcester, Essex, Suffolk, Bristol | same as hotspots |
| 26 | MI | Wayne, Oakland, Kent, Macomb, Genesee | same as hotspots |
| 27 | MN | Hennepin, St. Louis, Olmsted, Stearns, Dakota | same as hotspots |
| 28 | MS | Hinds, Harrison, DeSoto, Lee, Jackson | Hinds, Harrison, DeSoto, Lee, Lauderdale |
| 29 | MO | St. Louis, Jackson, St. Charles, Greene, Boone | St. Francois, Iron, Gentry, Shelby, Bollinger |
| 30 | MT | Yellowstone, Missoula, Gallatin, Lewis and Clark, Flathead | Gallatin, Yellowstone, Missoula, Lewis and Clark, Cascade |
| 31 | NE | Douglas, Lancaster, Sarpy, Hall, Buffalo | same as hotspots |
| 32 | NV | Clark, Washoe, Carson, Nye, Douglas | same as hotspots |
| 33 | NH | Hillsborough, Rockingham, Merrimack, Strafford, Grafton | same as hotspots |
| 34 | NJ | Bergen, Hudson, Monmouth, Ocean, Mercer | same as hotspots |

**Table S4:** Top five hotspots and superspreaders across the United States (NM–WY).

| **FIPS** | **State** | **Hotspots** | **Superspreaders** |
| --- | --- | --- | --- |
| 35 | NM | Bernalillo, Doña Ana, Santa Fe, San Juan, Eddy | Bernalillo, Santa Fe, Doña Ana, San Juan, Eddy |
| 36 | NY | New York, Kings, Suffolk, Westchester, Nassau | same as hotspots |
| 37 | NC | Wake, Mecklenburg, Guilford, Forsyth, Cumberland | same as hotspots |
| 38 | ND | Cass, Burleigh, Ward, Grand Forks, Morton | same as hotspots |
| 39 | OH | Franklin, Cuyahoga, Hamilton, Montgomery, Summit | same as hotspots |
| 40 | OK | Oklahoma, Tulsa, Comanche, Payne, Garfield | same as hotspots |
| 41 | OR | Multnomah, Washington, Marion, Lane, Deschutes | same as hotspots |
| 42 | PA | Philadelphia, Allegheny, Lancaster, Bucks, Montgomery | same as hotspots |
| 44 | RI | Providence, Kent, Washington, Newport, Bristol | same as hotspots |
| 45 | SC | Greenville, Richland, Charleston, Spartanburg, Horry | Richland, Charleston, Greenville, Lexington, Spartanburg |
| 46 | SD | Minnehaha, Pennington, Lincoln, Brookings, Brown | Minnehaha, Pennington, Lincoln, Brookings, Davison |
| 47 | TN | Davidson, Knox, Shelby, Rutherford, Hamilton | Davidson, Knox, Rutherford, Hamilton, Shelby |
| 48 | TX | Harris, Dallas, Tarrant, Bexar, Travis | same as hotspots |
| 49 | UT | Salt Lake, Utah, Davis, Weber, Washington | same as hotspots |
| 50 | VT | Chittenden, Washington, Windsor, Rutland, Franklin | same as hotspots |
| 51 | VA | Fairfax, Loudoun, Virginia Beach, Prince William, Chesterfield | same as hotspots |
| 53 | WA | King, Pierce, Snohomish, Spokane, Thurston | same as hotspots |
| 54 | WV | Kanawha, Monongalia, Cabell, Raleigh, Harrison | same as Hotspots |
| 55 | WI | Milwaukee, Dane, Waukesha, Brown, Kenosha | Milwaukee, Dane, Brown, Waukesha, Kenosha |
| 56 | WY | Natrona, Laramie, Albany, Campbell, Sweetwater | same as Hotspots |

**Table S5:** Stay-at-home orders across U.S. states during the COVID-19 pandemic, including effective dates and simulation periods (AL – NY).

| FIPS | State (Abbr.) | COVID-19 Lockdown Date (Stay-at-home Order)<br>Source: [50] | Simu. Days |
| --- | --- | --- | --- |
| 01 | Alabama (AL) | April 4 until April 30 | 95 |
| 02 | Alaska (AK) | March 28 until May 20 | 88 |
| 04 | Arizona (AZ) | March 31 until April 30 | 91 |
| 05 | Arkansas (AR) | No statewide stay-at-home order issued | 366 |
| 06 | California (CA) | March 19 | 79 |
| 08 | Colorado (CO) | March 26 until April 26 | 86 |
| 09 | Connecticut (CT) | March 23 until May 20 | 83 |
| 10 | Delaware (DE) | March 24 until May 15 | 84 |
| 11 | D.C. (DC) | April 1 until May 15 | 92 |
| 12 | Florida (FL) | April 3 until April 30 | 94 |
| 13 | Georgia (GA) | April 3 until April 30 | 94 |
| 15 | Hawaii (HI) | March 25 until May 31 | 85 |
| 16 | Idaho (ID) | March 25 until April 30 | 85 |
| 17 | Illinois (IL) | March 21 until April 30 | 81 |
| 18 | Indiana (IN) | March 24 until May 1 | 84 |
| 19 | Iowa (IA) | No statewide stay-at-home order issued | 366 |
| 20 | Kansas (KS) | March 30 until May 3 | 90 |
| 21 | Kentucky (KY) | March 26 | 86 |
| 22 | Louisiana (LA) | March 23 until May 15 | 83 |
| 23 | Maine (ME) | April 2 until May 31 | 93 |
| 24 | Maryland (MD) | March 30 until May 15 | 90 |
| 25 | Massachusetts (MA) | March 24 until May 4 | 84 |
| 26 | Michigan (MI) | March 24 until May 15 | 84 |
| 27 | Minnesota (MN) | March 27 until May 3 | 87 |
| 28 | Mississippi (MS) | April 3 until May 11 | 94 |
| 29 | Missouri (MO) | April 6 until May 3 | 97 |
| 30 | Montana (MT) | March 28 until April 10 | 88 |
| 31 | Nebraska (NE) | No statewide stay-at-home order issued | 366 |
| 32 | Nevada (NV) | April 1 until April 30 | 92 |
| 33 | New Hampshire (NH) | March 27 | 87 |
| 34 | New Jersey (NJ) | March 24 | 84 |
| 35 | New Mexico (NM) | March 24 until May 15 | 84 |
| 36 | New York (NY) | March 22 until May 15 | 82 |

**Table S6:**
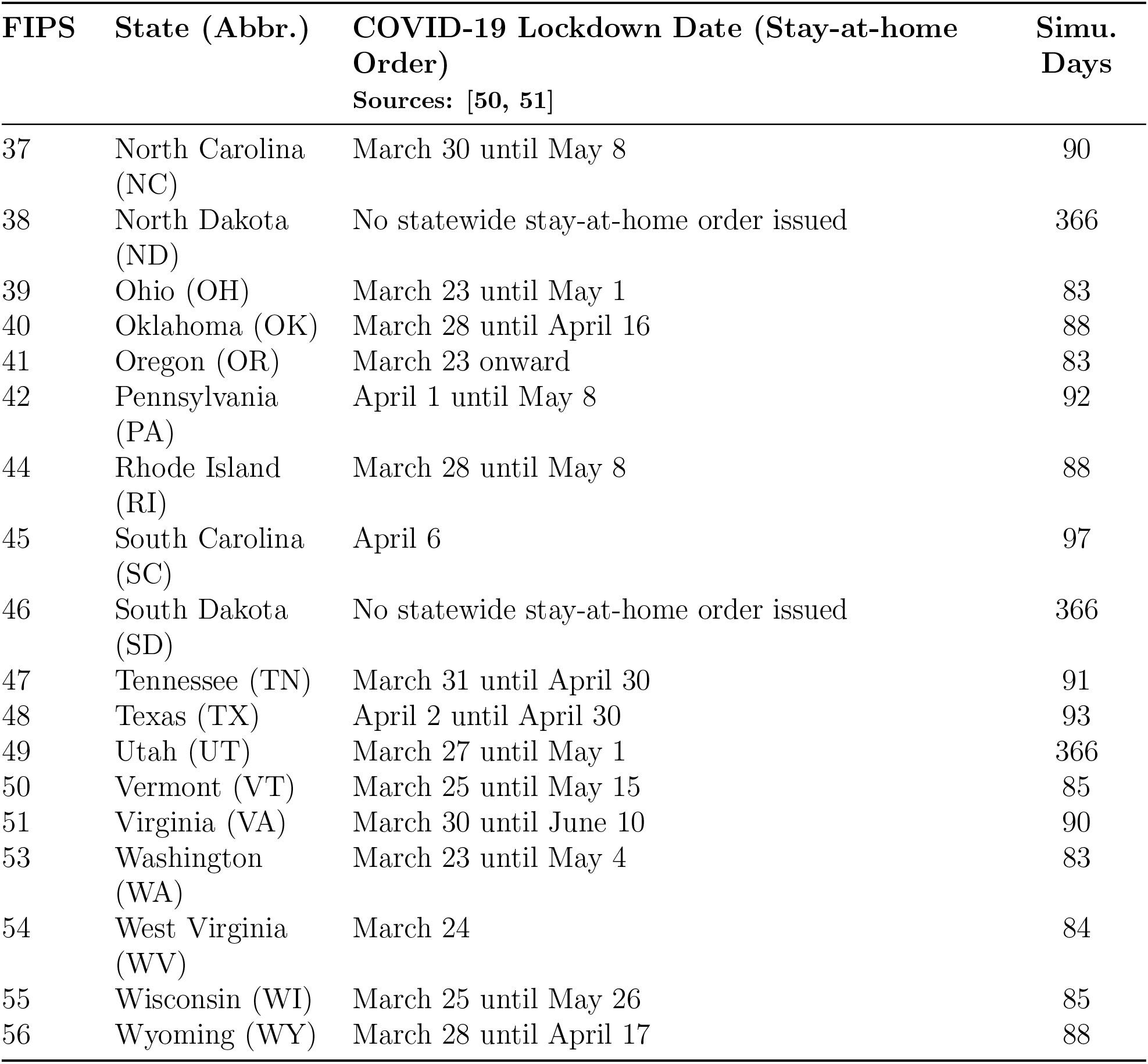
Stay-at-home orders across U.S. states during the COVID-19 pandemic, including effective dates and simulation periods (NC – WY).

